# Correlates of behavioral and emotional disorders among school-going adolescents in Uganda

**DOI:** 10.1101/2024.10.17.24315687

**Authors:** Max Bobholz, Julia Dickson-Gomez, Catherine Abbo, Arthur Kiconco, Abdul Shour, Simon Kasasa, Laura Cassidy, Ronald Anguzu

**Affiliations:** Medical College of Wisconsin, Milwaukee, US; Makerere University College of Health Sciences, Kampala, Uganda; Marshfield Clinic Health System, Marshfield, US; Department of Epidemiology and Biostatistics Makerere University School of Public Health Kampala, Uganda

**Keywords:** School mental health, adolescents, behavioral disorders, emotional disorders, Uganda

## Abstract

**Background:** Adolescence is a critical development transition period that increases vulnerability to poor mental health outcomes. Recent evidence suggests that 9.6% and 11.5% of adolescents in Uganda experienced behavioral and emotional disorders, respectively. We examined the factors associated with emotional and behavioral health outcomes among school-going adolescents in Uganda.

**Methods:** This cross-sectional study surveyed 1,953 students aged 10-24 enrolled in Central and Eastern Uganda secondary schools selected by stratified random sampling. Our outcome variables were (i) emotional and (ii) behavioral disorders that were measured using the Child and Adolescent Symptom Inventory-5 (CASI-5) diagnostic criteria outlined in the Diagnostic Statistical Manual-5 (DSM-5). Emotional disorders included major depressive disorder, generalized anxiety disorder, social anxiety disorder, and separation anxiety disorder. Attention deficit/hyperactivity disorder, conduct disorder, and oppositional defiant disorder were considered behavioral disorders. Covariates included socio-demographic, hardship-related experiences, and school-related characteristics. Modified Poisson and logistic regression models were appropriately run for the factors independently associated with respective outcomes. Prevalence ratios (PR), odds ratios (OR), and corresponding 95% confidence intervals (95%CI) were reported with p<0.05 considered significant.

**Results:** Participants’ mean age was 15.5 (SD=2.0) years; 54.7% were female, 5.7% had a behavioral disorder, and 17.4% had an emotional disorder. In the adjusted models, factors independently associated with higher odds of behavioral disorder were age (OR=1.2; 95%CI 1.1,1.4) and family history of mental illness (OR=1.9; 95%CI 1.2,3.3). Factors independently associated with a higher risk of emotional disorder were being female (PR=1.5; 95%CI 1.2,1.8), being enrolled in advanced education (PR=1.7; 95%CI 1.2,2.4), and attending private school (PR=1.4; 95%CI 1.1,1.8).

**Conclusions:** Behavioral and emotional disorders are prevalent among adolescents enrolled in secondary schools in Central and Eastern Uganda. Further inquiry using longitudinal designs is essential to understanding pathways for potential causality of the identified associations. School-based programs may consider routine screening for multi-level risk factors to improve the mental health of school-going adolescents.

## Background

Mental health conditions are a significant public health problem and are ranked the sixth leading cause of health loss and disability globally (1). An estimated 13.9% of the global population experiences mental disorders, with depression and anxiety disorders being the most common in all age groups. In adolescents (ages 10-19 years), one in seven experiences a mental disorder, which accounts for 13% of the global burden of disease in adolescents. Mental disorders, especially depression, have risen considerably in low, and middle-income countries (LMICs) (2). Depression, anxiety, and behavioral disorders are among the leading causes of illness and disability among adolescents, and suicide is the fourth leading cause of death among individuals aged 15 to 29 years. Depression alone accounts for 35% of the global burden of disease or disease-adjusted life years (DALYs), in addition to 700,000 annual deaths due to suicide (3,4). In Uganda, estimates show the prevalence of depression among children and adolescents at 23.6% (3). Mental health conditions can result from or lead to life difficulties, including poor relationships with family, friends, community, and problems at school and work.

Adolescence is a critical transition period during which biological and social development occurs (5). Developmental disruptions during this time can cause lasting developmental and health impacts on individuals (6–8). Since a substantial amount of an individual’s lifetime is spent in school settings during this period, the school environment plays a critical role in their social and emotional development. A safe and healthy school environment is essential for a child’s successful neurodevelopment and ability to thrive (9).

The burden of emotional and behavioral disorders among Uganda adolescents varies according to many factors, such as HIV status (10,11), trauma (12), and poverty (12,13). Recent studies using the Child and Adolescent Symptom Inventory-5 (CASI-5) among perinatally HIV-infected youth in Uganda have described a prevalence of emotional and behavioral disorders at 11.5% and 9.6%, respectively (14). In Uganda, Kinyanda and colleagues reported the risk factors for behavioral disorders as male and older (adolescent vs. child) (14). Another article utilizing similar methods for surveying symptoms reported a prevalence of ADHD at 6% among HIV-infected youth in Uganda. Few studies, however, have investigated the burden of emotional and behavioral disorders and the factors associated with these respective mental health disorders among school-going adolescents in Uganda (15–20). Similarly, little investigation has been done to understand the role of experiencing hardship and the individual’s school environment in mental health outcomes.

Screening for mental health disorders is an essential preventive mental health strategy aimed at reducing the occurrence and rising burden of mental disorders (21,22). Effective ways to survey adolescent mental and behavioral health have long been a focus of the scientific community; many test batteries have been developed to assess respondents’ burden of these outcomes. The CASI-5, published in 2013, was developed for children aged 5-18 as a behavior rating scale for conditions recognized by the Diagnostic Statistical Manual-5 (DSM-5) (23). The CASI-5 battery has been cross-culturally adapted for use in parts of Uganda (24).

Therefore, mental health in school settings should be a crucial focus of school administrators, public health practitioners, and policymakers. A recent systematic review showed that school-based mental health interventions can effectively improve mental health literacy and reduce stigma; heterogeneous methodologies and outcome assessments complicate this field of study (25). Coordinated comprehensive efforts of stakeholders such as school teachers and counselors are critical in addressing student mental health (26,27). Therefore, understanding the burden of emotional and behavioral health outcomes in secondary schools should inform stakeholder strategies to promote adolescent mental health and increase undetected mental health disorders while reducing their long-term negative health impact.

Although the burden of emotional and behavioral disorders among HIV-infected Ugandan youth has been explored, little investigation has been done to understand the role of experiencing hardship and the individual’s school environment in mental health outcomes. In the current study, we aimed to assess the factors associated with mental and behavioral health disorders among adolescents enrolled in secondary schools in central and eastern Uganda. We hypothesized that hardship experiences, demographic characteristics, and school features contribute to and could be associated with the prevalence of emotional and behavioral disorders among school-going adolescents in Uganda.

## Methods

### Study design, setting, and population

We used a cross-sectional study design to survey 1,972 adolescents aged 12 to 18 enrolled in eight secondary schools in Iganga district in Eastern Uganda and Mukono district in Central Uganda.

### Sampling strategy

We purposively selected Uganda’s two most populous regions, Central and Eastern. Iganga district schools (Eastern region) are predominantly rural, while Mukono district schools (Central region) are predominantly urban. For school sampling, we selected one school district from each region based on population and past academic performance. Using purposive, stratified random sampling, we selected eight secondary schools, four from each district. Each district had two government-funded and two private schools. We pre-visited the District Education Officers (DEO) for the list of all registered secondary schools in the respective districts. DEOs implement education laws and regulations according to government policies. We then created a list of schools and categorized them by their rural or urban status.

### Data collection and tools

Demographic surveys and CASI-5 questionnaires were administered by Psychiatric Clinical Officers (PCOs) who are healthcare providers trained and familiar with mental health diagnoses. The battery consists of more than one hundred questions about the frequency of a symptom’s presence, surveying 24 emotional and behavioral disorders. Each section of CASI-5 asks respondents about the symptoms of specific disorders. At the end of each section, a question is asked assessing how often the individual feels their symptoms impair social or behavioral function. Respondents were excluded from this analysis if they responded to less than half of the questions in at least one sub-section corresponding to emotional or behavioral disorders included in this analysis. For example, individuals who did not answer any questions on ADHD symptoms but completed all other pertinent questions were removed from the analysis. Less than 1% (n=19) of the study population was excluded, resulting in 1,953 remaining participants.

### Study measures

#### Dependent variables

The CASI-5 battery was used to screen selected study participants for the presence of symptoms of various mental health conditions. Responses to the CASI-5 survey are quantified by a Likert scale: 0 = Never, 1 = Sometimes, 2 = Often, 3 = Very Often. For this study, a response of at least 2 (Often or Very Often) indicated that the participant had the symptom. The DSM-5 guidelines were used to determine the presence of an emotional or behavioral disorder (28). Our composite categorical outcome variables (yes/no) were (i) the presence of an emotional disorder and (ii) the presence of a behavioral disorder. Emotional disorders used in this analysis include major depressive disorder, generalized anxiety disorder, social anxiety disorder, and separation anxiety disorder. Attention deficit/hyperactivity disorder, conduct disorder, and oppositional defiant disorder were included in behavioral disorder analyses.

#### Covariates

A demographic questionnaire was also administered alongside the CASI-5 battery. Demographic variables included sex (female/male), age (continuous), urbanity (rural/urban), and level of education [Advanced (A’) or Ordinary (O’) level]. Hardship variables were the nature of housing (permanent/ semi-permanent/hut), presence of domestic violence in the home (yes/no), family history of mental illness (yes/no), and orphanhood (at least one parental death/both parents living). School characteristics included sex status (mixed boys and girls/single sex) and school ownership (private/government). No manipulation was required for responses to the demographic survey as data did not require cleaning or recoding.

### Statistical Analysis

Descriptive statistics were obtained by computing frequencies and corresponding proportions of respondents who responded to the question and fit the emotional or behavioral disorder diagnostic criteria. In bivariate analysis, a chi-squared test of association was conducted between each categorical demographic characteristic and the presence of an emotional/behavioral disorder. Two-sample t-tests were used to assess mean age differences within each group. Further, bivariate logistic regression was performed for the presence of behavioral disorders, and modified Poisson regression (29) was used for analyzing associations between independent variables and the presence of an emotional disorder. Multivariable regression analyses were performed to understand the nature of associations while controlling for hardship experiences, demographic characteristics, and school features. We fitted the adjusted models with variables that were statistically significant at bivariate analysis and variables that were biologically plausible or potential confounders. The alpha threshold for statistical significance was set at 0.05.

## Results

### Participant demographics

Overall, among the study population of 1,953 school-going adolescents, their mean age was 15.47 years [standard deviation (SD)=1.99], and more were female (57.55%, n=1,124), urban dwellers (52.38%; n=1,023) and 12.35% (n=236) were enrolled in A’ level education. An overall proportion of 17.36% (n=340) reported symptoms suggestive of a mental health condition, and 5.71% (n=112) had behavioral disorders. Most respondents (59.04%; n=1,153) attended private schools, and 89.71% (n=1,752) studied in mixed-gender schools. In terms of hardship experiences of respondents, 14.03% (n=263) reported witnessing domestic violence in the home setting, 18.82% had a family history of mental illness, and 17.10% (n=332) were single or two-parent orphans. Few respondents (1.48%; n=28) reported living in hut-style, grass-thatched homes, 82.40% (n=1,559) lived in permanent residences, and 16.12% of respondents (n=305) lived in semi-permanent homes (**Table 1**).

**Table 1.** Bivariate tests of simple association (chi-squared) among demographic characteristics, hardship experiences, school features, and the presence of emotional or behavioral disorders among Ugandan school-going adolescents.

|  |  | Emotional disorder |  |  | Behavioral disorder |  |  |
| --- | --- | --- | --- | --- | --- | --- | --- |
| Characteristic | Totals<br>n (%) | Yes<br>n (%) | No<br>n (%) | $\chi^2$ | Yes<br>n (%) | No<br>n (%) | $\chi^2$ |
| **Sex** |  |  |  |  |  |  |  |
| Male | 829 (42.45) | 113 (13.61) | 717 (86.39) | 14.121*** | 48 (5.76) | 785 (94.24) | 0.006 |
| Female | 1,124 (57.55) | 227 (20.12) | 901 (79.88) |  | 64 (5.68) | 1,063 (94.32) |  |
| **Age [mean (sd)] †** | 15.472 (1.99) | 15.400 | 15.486 | p=0.224 | 15.938 | 15.441 | p=0.005** |
| **Urbanity** |  |  |  |  |  |  |  |
| Rural | 930 (47.62) | 138 (14.81) | 794 (85.19) | 8.109** | 47 (5.03) | 888 (94.97) | 1.569 |
| Urban | 1,023 (52.38) | 202 (19.69) | 824 (80.31) |  | 65 (6.34) | 960 (93.66) |  |
| **Level of education enrolled in** |  |  |  |  |  |  |  |
| Advanced level | 236 (12.35) | 49 (20.76) | 187 (79.24) | 2.005 | 14 (5.91) | 223 (94.09) | 0.015 |
| Ordinary level | 1,675 (87.65) | 286 (17.02) | 1,394 (82.98) |  | 96 (5.71) | 1,585 (94.29) |  |
| **Nature of housing** |  |  |  |  |  |  |  |
| Permanent | 1,559 (82.40) | 257 (16.45) | 1,305 (83.55) | 8.136* | 85 (5.43) | 1,481 (94.57) | 4.108 |
| Semi-permanent | 305 (16.12) | 68 (22.15) | 239 (77.85) |  | 18 (5.90) | 287 (94.10) |  |
| Hut | 28 (1.48) | 8 (28.57) | 20 (71.43) |  | 4 (14.29) | 24 (85.71) |  |
| **Domestic violence witness** |  |  |  |  |  |  |  |
| Yes | 263 (14.03) | 58 (22.05) | 205 (77.95) | 4.738* | 21 (7.95) | 243 (92.05) | 3.106 |
| No | 1,612 (85.97) | 268 (16.57) | 1,349 (83.43) |  | 85 (5.26) | 1,532 (94.74) |  |
| **Family history of mental illness** |  |  |  |  |  |  |  |
| Yes | 328 (18.82) | 73 (22.26) | 255 (77.74) | 5.322* | 31 (9.42) | 298 (90.58) | 11.152*** |
| No | 1,415 (81.18) | 239 (16.84) | 1,180 (83.16) |  | 67 (4.72) | 1,352 (95.28) |  |
| **Orphan** |  |  |  |  |  |  |  |
| Yes | 332 (17.10) | 62 (18.62) | 271 (81.38) | 0.401 | 16 (4.78) | 319 (95.22) | 0.703 |

\* <0.05; \*\*<0.01; \*\*\*<0.001

|  |  |  |  |  |  |  |  |
| --- | --- | --- | --- | --- | --- | --- | --- |
| No | 1,610 (82.90) | 277 (17.17) | 1,336 (82.83) |  | 96 (5.95) | 1,518 (94.05) |  |
| **School sex status** |  |  |  |  |  |  |  |
| Mixed | 1,752 (89.71) | 310 (17.64) | 1,447 (82.36) | 0.929 | 101 (5.75) | 1,657 (94.25) | 0.030 |
| Single-sex | 201 (10.29) | 30 (14.93) | 171 (85.07) |  | 11 (5.45) | 191 (94.55) |  |
| **School ownership** |  |  |  |  |  |  |  |
| Private | 1,153 (59.04) | 168 (20.95) | 984 (79.05) | 12.153*** | 45 (5.62) | 755 (94.38) | 0.020 |
| Government | 800 (40.96) | 172 (14.88) | 634 (85.12) |  | 67 (5.78) | 1,093 (94.22) |  |
| **Total** | 1,953 (100) | 340 (17.36) | 1,618 (82.64) | -- | 112 (5.71) | 1,848 (94.29) | -- |

† : a t-test was performed to assess differences in mean age

### Prevalence of behavioral disorders

The prevalence of the behavioral disorders ranged from 1.89% (conduct disorder) to 2.96% (opposition defiant disorder). The lowest prevalence of emotional disorders was 0.20% for major depressive disorder, with the highest prevalence of 14.20% for separation anxiety disorder. Low depression prevalence was reinforced by applying the symptom severity T-score approach (23), observing that only mild depression was present; no individuals met the threshold for moderate or severe depression with this method. The prevalence of individual emotional and behavioral disorders can be found in **Supplemental Table 1**.

### Prevalence and correlates of emotional disorders

In the bivariate regression analyses, females (PR=1.48; 95%CI 1.20,1.82) and attending private school (PR=1.41; 95%CI 1.16,1.71) had 1.5 and 1.4 times the prevalence of emotional disorders when compared to male students and attending public schools, respectively. In other significant bivariate regression analyses, the prevalence of emotional disorders was significantly higher among urban-dwelling students (PR=1.33; 95%CI 1.09,1.62), those living in semi-permanent homes (PR=1.35; 95%CI 1.06,1.71), those who witnessed domestic violence (PR=1.33; 95%CI 1.03,1.71), and those who had a family history of mental illness (PR=1.32; 95%CI 1.05,1.67). In the adjusted regression model, factors independently associated with the presence of an emotional disorder were being female (adj. PR=1.46; 95%CI 1.15,1.84), compared to male students, and attending private school (adj. PR=1.40; 95%CI 1.12,1.76), compared to attending public school (**Table 2**; **Figure 1**).

**Table 2.** Logistic and modified Poisson regression models for assessing association of demographic characteristics, hardship experiences, school features and the presence of emotional or behavioral disorders among Ugandan school-going adolescents.

| Characteristic | Emotional disorder |  |  |  | Behavioral Disorder |  |  |  |
| --- | --- | --- | --- | --- | --- | --- | --- | --- |
|  | PR | 95% CI | APR | 95% CI | OR | 95% CI | AOR | 95% CI |
| **Sex** |  |  |  |  |  |  |  |  |
| Male | Ref | -- | Ref | -- | Ref | -- | Ref | -- |
| Female | 1.478*** | 1.20-1.82 | 1.457** | 1.15-1.84 | 0.985 | 0.67-1.45 | 1.156 | 0.72-1.85 |
| **Age [mean (sd)]** | 0.982 | 0.93-1.03 | 0.966 | 0.91-1.03 | 1.126* | 1.03-1.23 | 1.205** | 1.05-1.38 |
| **Urbanity** |  |  |  |  |  |  |  |  |
| Rural | Ref | -- | Ref | -- | Ref | -- | Ref | -- |
| Urban | 1.330** | 1.09-1.62 | 1.216 | 0.96-1.54 | 1.279 | 0.87-1.88 | 1.224 | 0.76-1.98 |
| **Level of education enrolled in** |  |  |  |  |  |  |  |  |
| Advanced level | 1.220 | 0.93-1.60 | 1.690** | 1.18-2.42 | 1.036 | 0.58-1.85 | 0.670 | 0.31-1.44 |
| Ordinary level | Ref | -- | Ref | -- | Ref | -- | Ref | -- |
| **Nature of housing** |  |  |  |  |  |  |  |  |
| Permanent | Ref | -- | Ref | -- | Ref | -- | Ref | -- |
| Semi-permanent | 1.346* | 1.06-1.71 | 1.260 | 0.98-1.63 | 1.093 | 0.65-7.85 | 1.049 | 0.60-1.83 |
| Hut | 1.737 | 0.96-3.15 | 1.306 | 0.67-2.56 | 2.904 | 0.99-8.56 | 1.523 | 0.41-5.70 |

\* <0.05; \*\*<0.01; \*\*\*<0.001

|  |  |  |  |  |  |  |  |  |
| --- | --- | --- | --- | --- | --- | --- | --- | --- |
| **Domestic violence witness** |  |  |  |  |  |  |  |  |
| Yes | 1.331* | 1.03-1.71 | 1.230 | 0.93-1.63 | 1.558 | 0.95-2.56 | 1.476 | 0.84-2.59 |
| No | Ref | -- | Ref | -- | Ref | -- | Ref | -- |
| **Family history of mental illness** |  |  |  |  |  |  |  |  |
| Yes | 1.321* | 1.05-1.67 | 1.163 | 0.90-1.50 | 2.099** | 1.35-3.27 | 1.994** | 1.22-3.26 |
| No | Ref | -- | Ref | -- | Ref | -- | Ref | -- |
| **Orphan** |  |  |  |  |  |  |  |  |
| Yes | 1.084 | 0.85-1.39 | 1.028 | 0.79-1.39 | 0.793 | 0.46-1.36 | 0.700 | 0.39-1.27 |
| No | Ref | -- | Ref | -- | Ref | -- | Ref | -- |
| **School sex status** |  |  |  |  |  |  |  |  |
| Mixed | 1.182 | 0.84-1.67 | 0.935 | 0.60-1.45 | 1.058 | 0.56-2.01 | 0.792 | 0.34-1.86 |
| Single-sex | Ref | -- | Ref | -- | Ref | -- | Ref | -- |
| **School ownership** |  |  |  |  |  |  |  |  |
| Private | 1.408** | 1.16-1.71 | 1.403** | 1.12-1.76 | 0.972 | 0.66-1.43 | 0.881 | 0.55-1.40 |
| Government | Ref | -- | Ref | -- | Ref | -- | Ref | -- |

**Abbreviations:** PR = Prevalence Ratio; APR = Adjusted Prevalence Ratio; OR = Odds Ratio; AOR = Adjusted Odds Ratio; Ref = Referent

**Figure 1.**
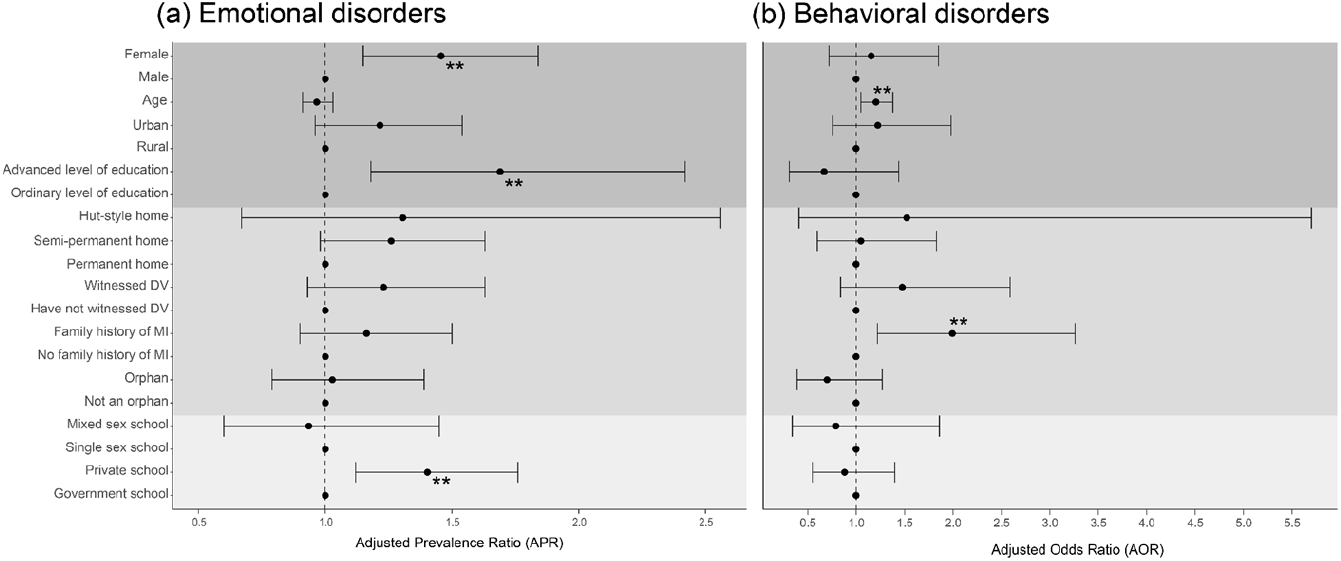
Logistic regression modeling for (a) emotional and (b) behavioral disorders adjusted for demographic factors (dark grey), hardship experiences (grey), and school features (light grey) among Ugandan school-going adolescents. * = p<0.05; ** = p<0.01; *** = p<0.001

### Prevalence and correlates of behavioral disorders

In the unadjusted regression analyses, age (OR=1.13; 95%CI 1.03,1.23) and having a family history of mental illness (OR=2.10; 95%CI 1.35,3.27) were associated with having a behavioral disorder. These associations persisted in the adjusted model, as being one year older (adj. OR=1.21; 95%CI 1.05,1.38) and having a family history of mental illness (adj. OR=1.99; 95%CI 1.22,3.26) had 1.2 and 2.0 times the prevalence of behavioral disorders as compared to younger individuals without mental illness in the family (Table 2; Figure 1).

## Discussion

This study explored the factors associated with emotional and behavioral disorders among school-going adolescents in Uganda. We revealed two key findings. First, being female and attending private schools were independently associated with emotional disorders. Secondly, older age and having a family history of mental illness were independently associated with behavioral disorders among adolescents attending secondary schools in Central and Eastern Uganda.

The prevalence of emotional disorders (depression, generalized anxiety, PTSD, social anxiety, and separation anxiety) among this sample of Ugandan adolescents was slightly higher than the values reported in another study from Uganda using the CASI-5 battery (14). The same study also reported a prevalence of behavioral disorders at 9.6%, slightly higher than the prevalence reported in this study. However, the combined prevalence of any emotional or behavioral disorder in this study’s sample (20.2%) is consistent with a systematic review of mental disorders in Uganda (30). The cross-cultural adaptability of the CASI-5 tool has been assessed and documented (24).

The prevalence of psychiatric disorders among school-going adolescents in India was greater than what was observed in this study, with the significant difference being a higher prevalence of depression (31). A recent systematic review of mental health problems among adolescents in Sub-Saharan Africa reported a similar prevalence of mental health problems, except for a much larger prevalence of depression (32). This discrepancy should caution against the generalizability of this study’s findings to depression outcomes. By using an alternative approach to categorizing depression in this population (specifically the Symptom Severity T-score approach (23)), we confirmed that this study population had no respondents who fit the threshold for moderate or severe depression.

Our study found that female students were at increased risk for having emotional disorders; however, sex was not significantly associated with having a behavioral disorder. Significant differences in mental and behavioral health outcomes among Ugandan youth according to sex have been found (14,33,34). Our study revealed twice as many female adolescent students having emotional disorders than their male counterparts. Prior studies consistently reveal that a higher burden of disorders such as depression among females may linked to biological or socio-cultural experiences (35–37).

School-based interventions have become attractive opportunities to improve mental health outcomes among adolescents who attend school (38–40). Our study’s findings suggest that further attention is required for Ugandan private schools to promote mental health. A school culture beyond academic responsibility and fostering positive relationships with teachers has been highlighted as potentially crucial for school-based mental health interventions (41). This study also highlights potential populations to focus interventions on, including, but not limited to, those attending private schools, those with a family history of mental illness, and female students in Uganda. Each of these factors has been shown to independently increase one’s risk of having an emotional or behavioral disorder in this study population. Prior research demonstrated that economic and family interventions reduced absenteeism (general and sickness-related) among school-going adolescent girls (42). However, this intervention did not improve behavior and grade repetition among schooling adolescent girls in Southern Uganda.

As a result of the cross-sectional study design, this study can only highlight associations with emotional or behavioral disorders; inferences of causality cannot be made with this study’s findings alone. The length of the CASI-5 test battery could have introduced potential biases or misrepresentations in this dataset. Respondents answered 202 questions about symptoms they may be experiencing, possible impairment resulting from these symptoms, and demographic details. Suboptimal effort and fatigue while testing is a concern among neuropsychological scientists assessing mental health outcomes in children (43). Although there was potential for testing fatigue biasing the dataset, we attempted to address this by removing observations where individuals responded to less than half of the survey. Respondents self-reported symptom frequency and symptom impairment, which may have contributed to slight misrepresentations of true emotional and behavioral health burdens on this population. Major depressive disorder was minimally present within this study population, which differs from other studies; this observation should be explored further to assess whether there are socio-cultural, economic, or environmental explanations beyond tool validity with this population. The rarity of some key independent variable outcomes may have limited this study. For example, very few respondents reported living in huts. In bivariate and adjusted models, living in a hut carried a large effect size, but the rarity of this outcome reduced the findings’ statistical confidence. This, however, highlights an area of future investigation.

## Conclusions

Being female, having an advanced level of education, and attending a private school are independently associated with the presence of an emotional disorder, while age and having a family history of mental illness were independently associated with having a behavioral disorder. These findings should inform Ugandan officials and researchers focusing on improving adolescent emotional and behavioral health outcomes. Additionally, we have addressed potential areas for school-based interventions and highlighted potentially at-risk populations. Further research is required to assess the causality of such results, understand what key experiences may have contributed to poor emotional or behavioral health, and the contributions of less stable housing and other hardship experiences in mental health.

## Data Availability

The deidentified dataset used for cleaning and analysis is available for download.

## List of abbreviations

CASI-5: Child and Adolescent Symptom Inventory-5
PR: prevalence ratio
OR: odds ratio
APR: adjusted prevalence ratio
AOR: adjusted odds ratio

## Supplemental Information

**Supplemental Table 1**. Detailed summary statistics for each emotional and behavioral disorder included in this analysis among Ugandan school-going adolescents.

## Declarations

## Acknowledgments

We also thank the secondary students’ surveys for their support and cooperation.

## Availability of data and material

All relevant data are within the manuscript.

## Authors’ contributions

Conceptualization: Catherine Abbo, Max Bobholz, Ronald Anguzu, Methodology: Catherine Abbo, Max Bobholz, Ronald Anguzu, Laura Cassidy, Writing – original draft: Max Bobholz, Writing – review & editing: Max Bobholz, Julia Dickson-Gomez, Catherine Abbo, Arthur Kiconco, Abdul R. Shour, Simon Kasasa, Laura Cassidy, Ronald Anguzu

## Consent for publication

Not applicable.

## Ethics approval and consent to participate

Ethical approval was obtained from the School of Medicine Research Ethics Committee (SOMREC). Permission to conduct the study was also obtained from the Uganda National Council for Science and Technology (UNCST). All respondents provided informed consent to participate in the study.

## References

1. Institute for Health Metrics and Evaluation [Internet]. [cited 2024 Jul 21]. Mental health. Available from: https://www.healthdata.org/research-analysis/health-risks-issues/mental-health

2. Okello ES, Neema S. Explanatory models and help-seeking behavior: Pathways to psychiatric care among patients admitted for depression in Mulago hospital, Kampala, Uganda. Qual Health Res. 2007 Jan;17(1):14–25.

3. Kaggwa MM, Najjuka SM, Bongomin F, Mamun MA, Griffiths MD. Prevalence of depression in Uganda: A systematic review and meta-analysis. PLOS ONE. 2022 Oct 20;17(10):e0276552.

4. Depressive disorder (depression) [Internet]. [cited 2024 Jul 21]. Available from: https://www.who.int/news-room/fact-sheets/detail/depression

5. Dahl RE, Allen NB, Wilbrecht L, Suleiman AB. Importance of investing in adolescence from a developmental science perspective. Nature. 2018 Feb;554(7693):441–50.

6. Chaby LE, Zhang L, Liberzon I. The effects of stress in early life and adolescence on posttraumatic stress disorder, depression, and anxiety symptomatology in adulthood. Curr Opin Behav Sci. 2017 Apr 1;14:86–93.

7. Guessoum SB, Lachal J, Radjack R, Carretier E, Minassian S, Benoit L, et al. Adolescent psychiatric disorders during the COVID-19 pandemic and lockdown. Psychiatry Res. 2020 Sep 1;291:113264.

8. Larsen B, Luna B. Adolescence as a neurobiological critical period for the development of higher-order cognition. Neurosci Biobehav Rev. 2018 Nov 1;94:179–95.

9. Themane M, Osher D. Schools as enabling environments. South Afr J Educ. 2014 Nov;34(4):1–6.

10. Mpango RS, Kinyanda E, Rukundo GZ, Levin J, Gadow KD, Patel V. Prevalence and correlates for ADHD and relation with social and academic functioning among children and adolescents with HIV/AIDS in Uganda. BMC Psychiatry. 2017 Sep 22;17(1):336.

11. Mpango RS, Rukundo GZ, Muyingo SK, Gadow KD, Patel V, Kinyanda E. Prevalence, correlates for early neurological disorders and association with functioning among children and adolescents with HIV/AIDS in Uganda. BMC Psychiatry. 2019 Jan 21;19(1):34.

12. Onyut LP, Neuner F, Ertl V, Schauer E, Odenwald M, Elbert T. Trauma, poverty and mental health among Somali and Rwandese refugees living in an African refugee settlement – an epidemiological study. Confl Health. 2009 May 26;3(1):6.

13. Karimli L, Ssewamala FM, Neilands TB. The impact of poverty-reduction intervention on child mental health mediated by family relations: Findings from a cluster-randomized trial in Uganda. Soc Sci Med. 2023 Sep 1;332:116102.

14. Kinyanda E, Salisbury TT, Levin J, Nakasujja N, Mpango RS, Abbo C, et al. Rates, types and co-occurrence of emotional and behavioural disorders among perinatally HIV-infected youth in Uganda: the CHAKA study. Soc Psychiatry Psychiatr Epidemiol. 2019 Apr 1;54(4):415–25.

15. Nalugya-Sserunjogi J, Rukundo GZ, Ovuga E, Kiwuwa SM, Musisi S, Nakimuli-Mpungu E. Prevalence and factors associated with depression symptoms among school-going adolescents in Central Uganda. Child Adolesc Psychiatry Ment Health. 2016;10:39.

16. Okello J, Nakimuli-Mpungu E, Musisi S, Broekaert E, Derluyn I. The Association between Attachment and Mental Health Symptoms among School-Going Adolescents in Northern Uganda: The Moderating Role of War-Related Trauma. PLoS ONE. 2014 Mar 10;9(3):e88494.

17. Nabunya P, Damulira C, Byansi W, Muwanga J, Bahar OS, Namuwonge F, et al. Prevalence and correlates of depressive symptoms among high school adolescent girls in southern Uganda. BMC Public Health. 2020 Nov 25;20(1):1792.

18. Rudatsikira E, Muula AS, Siziya S, Twa-Twa J. Suicidal ideation and associated factors among school-going adolescents in rural Uganda. BMC Psychiatry. 2007 Nov 23;7(1):67.

19. Ssewamala FM, Brathwaite R, Neilands TB. Economic Empowerment, HIV Risk Behavior, and Mental Health Among School-Going Adolescent Girls in Uganda: Longitudinal Cluster-Randomized Controlled Trial, 2017–2022. Am J Public Health. 2023 Mar;113(3):306–15.

20. Byansi W, Ssewamala FM, Neilands TB, Mwebembezi A, Nakigozi G. Patterns of and Factors Associated With Mental Health Service Utilization Among School-Going Adolescent Girls in Southwestern Uganda: A Latent Class Analysis. J Adolesc Health Off Publ Soc Adolesc Med. 2023 May;72(5S):S24–32.

21. Arango C, Díaz-Caneja CM, McGorry PD, Rapoport J, Sommer IE, Vorstman JA, et al. Preventive strategies for mental health. Lancet Psychiatry. 2018 Jul;5(7):591–604.

22. Prevention and promotion in mental health. World Health Organization: Department of Mental Health and Substance Dependence; 2002.

23. Checkmate Plus Site [Internet]. [cited 2024 Mar 5]. Child & Adolescent Symptom Inventory-5. Available from: https://www.checkmateplus.com/casi-5

24. Mpango RS, Kinyanda E, Rukundo GZ, Gadow KD, Patel V. Cross-cultural adaptation of the Child and Adolescent Symptom Inventory-5 (CASI-5) for use in central and south-western Uganda: the CHAKA project. Trop Doct. 2017 Oct 1;47(4):347–54.

25. Ma KKY, Anderson JK, Burn AM. Review: School-based interventions to improve mental health literacy and reduce mental health stigma – a systematic review. Child Adolesc Ment Health. 2023;28(2):230–40.

26. Beames JR, Johnston L, O’Dea B, Torok M, Boydell K, Christensen H, et al. Addressing the mental health of school students: Perspectives of secondary school teachers and counselors. Int J Sch Educ Psychol. 2022 Jan 2;10(1):128–43.

27. Collins TP. Addressing Mental Health Needs in Our Schools: Supporting the Role of School Counselors. Prof Couns. 2014;4(5):413–6.

28. American Psychiatric Association. Diagnostic and Statistical Manual of Mental Disorders. 5th ed. 2013. 947 p.

29. Zou G. A modified poisson regression approach to prospective studies with binary data. Am J Epidemiol. 2004 Apr 1;159(7):702–6.

30. Opio JN, Munn Z, Aromataris E. Prevalence of Mental Disorders in Uganda: a Systematic Review and Meta-Analysis. Psychiatr Q. 2022 Mar 1;93(1):199–226.

31. Shafi M, Younis N, Rasool U, Younis S, Rather YH, Lone BB, et al. Prevalence of psychiatric morbidity among school-going adolescents in the age group of 13–19 years. Middle East Curr Psychiatry. 2023 Aug 7;30(1):64.

32. Hunduma G, Dessie Y, Geda B, Yadeta TA, Deyessa N. Common mental health problems among adolescents in sub-Saharan Africa: A systematic review and meta-analysis. J Child Adolesc Ment Health. 2021 Sep 2;33(1–3):90–110.

33. Cohen F, Seff I, Ssewamala F, Opobo T, Stark L. Intimate Partner Violence and Mental Health: Sex-Disaggregated Associations Among Adolescents and Young Adults in Uganda. J Interpers Violence. 2022 Mar 1;37(5–6):2399–415.

34. Abbo C, Kinyanda E, Kizza RB, Levin J, Ndyanabangi S, Stein DJ. Prevalence, comorbidity and predictors of anxiety disorders in children and adolescents in rural north-eastern Uganda. Child Adolesc Psychiatry Ment Health. 2013 Jul 10;7(1):21.

35. Anguzu R, Akun P, Katairo T, Abbo C, Ningwa A, Ogwang R, et al. Household poverty, schooling, stigma and quality of life in adolescents with epilepsy in rural Uganda. Epilepsy Behav. 2021 Jan 1;114:107584.

36. Allen JP, Pettit C, Costello MA, Hunt GL, Stern JA. A social-development model of the evolution of depressive symptoms from age 13 to 30. Dev Psychopathol. 2024 Feb;36(1):280–90.

37. Chaplin TM, Gillham JE, Seligman MEP. Gender, Anxiety, and Depressive Symptoms: A Longitudinal Study of Early Adolescents. J Early Adolesc. 2009 Apr 1;29(2):307–27.

38. Paulus FW, Ohmann S, Popow C. Practitioner Review: School-based interventions in child mental health. J Child Psychol Psychiatry. 2016;57(12):1337–59.

39. Schachter HM, Girardi A, Ly M, Lacroix D, Lumb AB, van Berkom J, et al. Effects of school-based interventions on mental health stigmatization: a systematic review. Child Adolesc Psychiatry Ment Health. 2008 Jul 21;2(1):18.

40. Shoshani A, Steinmetz S. Positive Psychology at School: A School-Based Intervention to Promote Adolescents’ Mental Health and Well-Being. J Happiness Stud. 2014 Dec 1;15(6):1289–311.

41. Carlson C, Namy S, Nakuti J, Mufson L, Ikenberg C, Musoni O, et al. Student, teacher, and caregiver perceptions on implementing mental health interventions in Ugandan schools. Implement Res Pract. 2021 Jan 1;2:26334895211051290.

42. Brathwaite R, Namuwonge F, Magorokosho N, Tutlam N, Neilands TB, Namirembe R, et al. Impact of Economic and Family Intervention on Adolescent Girls’ Education Performance, School Absenteeism, and Behavior in School: The Suubi4Her Study. J Adolesc Health. 2024 Feb 1;74(2):340–9.

43. DeRight J, Carone DA. Assessment of effort in children: A systematic review. Child Neuropsychol. 2015 Jan 2;21(1):1–24.

